# Structure-function multilayer network integration and cognition in multiple sclerosis

**DOI:** 10.1101/2025.03.31.25324960

**Authors:** Lucas C. Breedt, Giuseppe Pontillo, Fernando A.N. Santos, Chris Vriend, Ferran Prados, Alle Meije Wink, Alvino Bisecco, Alessandro Cagol, Massimiliano Calabrese, Marco Castellaro, Sara Collorone, Rosa Cortese, Nicola De Stefano, Christian Enzinger, Massimo Filippi, Michael A. Foster, Antonio Gallo, Gabriel Gonzalez-Escamilla, Cristina Granziera, Sergiu Groppa, Einar A. Høgestøl, Sara Llufriu, Eloy Martinez-Heras, Elisabeth Solana, Silvia Messina, Marcello Moccia, Gro O. Nygaard, Jacqueline Palace, Daniela Pinter, Mara A. Rocca, Ahmed Toosy, Paola Valsasina, Olga Ciccarelli, Eva M. Strijbis, Frederik Barkhof, Menno M. Schoonheim, Linda Douw, MAGNIMS study group

**Affiliations:** Department of Anatomy and Neurosciences, Amsterdam Neuroscience, Amsterdam UMC, Vrije Universiteit Amsterdam, Amsterdam, The Netherlands; MS Center Amsterdam, Amsterdam Neuroscience, Amsterdam UMC, Vrije Universiteit Amsterdam, Amsterdam, The Netherlands; Queen Square Multiple Sclerosis Centre, Department of Neuroinflammation, UCL Queen Square Institute of Neurology, University College London, London, United Kingdom; Department of Radiology and Nuclear Medicine, MS Center Amsterdam, Amsterdam Neuroscience, Amsterdam UMC, Vrije Universiteit Amsterdam, Amsterdam, The Netherlands; Departments of Advanced Biomedical Sciences and Electrical Engineering and Information Technology, University of Naples “Federico II”, Naples, Italy; Dutch Institute for Emergent Phenomena (DIEP), Institute for Advanced Studies, University of Amsterdam, Amsterdam, The Netherlands; Department of Psychiatry, Amsterdam Neuroscience, Amsterdam UMC, Vrije Universiteit Amsterdam, Amsterdam, The Netherlands; Centre for Medical Image Computing, Department of Medical Physics and Biomedical Engineering, University College London, London, United Kingdom; E-Health Center, Universitat Oberta de Catalunya, Barcelona, Spain; Department of Advanced Medical and Surgical Sciences, University of Campania “Luigi Vanvitelli”, Naples, Italy; Translational Imaging in Neurology (ThINK) Basel, Department of Biomedical Engineering, Faculty of Medicine, University Hospital Basel and University of Basel, Basel, Switzerland; Department of Neurology, University Hospital Basel, Switzerland; Research Center for Clinical Neuroimmunology and Neuroscience Basel (RC2NB), University Hospital Basel and University of Basel, Basel, Switzerland; Department of Neurosciences, Biomedicine and Movement Sciences, University of Verona, Verona, Italy; Department of Information Engineering, University of Padova, Padova, Italy; Department of Medicine, Surgery and Neuroscience, University of Siena, Siena, Italy; Department of Neurology, Medical University of Graz, Graz, Austria; Movement Disorders, Neurostimulation and Neuroimaging, University Medicine Mainz, Mainz, Germany; Department of Neurology, Oslo University Hospital, Oslo, Norway; Department of Psychology, University of Oslo, Oslo, Norway; Neuroimmunology and Multiple Sclerosis Unit and Laboratory of Advanced Imaging in Neuroimmunological Diseases (ImaginEM), Hospital Clinic and Institut d’Investigacions Biomèdiques August Pi i Sunyer (IDIBAPS), University of Barcelona, Barcelona, Spain; Nuffield Department of Clinical Neurosciences, University of Oxford, Oxford, United Kingdom; Department of Molecular Medicine and Medical Biotechnology, University of Naples “Federico II”, Naples, Italy; Neuroimaging Research Unit, Division of Neuroscience, IRCCS San Raffaele Scientific Institute, Milan, Italy; Neurology Unit, IRCCS San Raffaele Scientific Institute, Milan, Italy; Neurorehabilitation Unit, IRCCS San Raffaele Scientific Institute, Milan, Italy; Neurophysiology Service, IRCCS San Raffaele Scientific Institute, Milan, Italy; Vita-Salute San Raffaele University, Milan, Italy; Department of Neurology, MS Center Amsterdam, Amsterdam Neuroscience, Amsterdam UMC, Vrije Universiteit Amsterdam, Amsterdam, The Netherlands; Centre for Medical Image Computing, Department of Computer Science, University College London, London, United Kingdom; Dementia Research Centre, UCL Queen Square Institute of Neurology, University College London, London, United Kingdom; Institute of Clinical Medicine, University of Oslo, Oslo, Norway; Dipartimento di Scienze della Salute, Università degli Studi di Genova

## Abstract

People with multiple sclerosis (MS) often present with cognitive deficits that cannot fully be attributed to focal brain alterations. Whole-brain network changes show stronger relations, but MS network insights have mostly focused on either structural or functional (single-layer) networks, while recent work has shown the importance of multilayer frontoparietal network integration for cognition. Here, we explored the cognitive relevance of multilayer integration of the frontoparietal network in relapsing-remitting MS (n=780) using diffusion and resting-state functional MRI. Cognitive relations were first assessed for nodal multilayer eigenvector centrality, averaged over frontoparietal network nodes as a measure of integration; and post-hoc for mean eccentricity for both single-layers and the multilayer.

Higher multilayer frontoparietal network centrality was associated with worse SDMT performance (β = −0.117, p = 0.005). Mean eccentricity of single-layer diffusion (β = −0.123, p < 0.001) and multilayer networks (β = 0.085, p = 0.018) were associated with cognition. However, results could not be replicated using a different anatomical parcellation.

This study showed that cognition in MS is related to multilayer network parameters. Nevertheless, correlations were weak and atlas-specific, suggesting that a binary structure-function multilayer network approach is not particularly relevant as a correlate of cognition in MS.

## 1. Introduction

People with multiple sclerosis (MS) often suffer from cognitive impairment [1], impacting their quality of life [2, 3]. Although the pathophysiology of MS centers around demyelination and hence the presence of focal white- and grey matter lesions, lesion load does not necessarily explain cognitive dysfunction. This discrepancy is known as the clinico-radiological paradox [4] and has led to the search for more advanced imaging markers to better understand clinical symptoms and progression in MS.

In light of this, it has become common practice to view the brain as a complex network through the lens of graph theory [5, 6]. In this framework, brain regions are represented as nodes, while the interactions between brain regions form the connections or edges between the nodes. These connections can be derived from structural white matter pathways obtained through diffusion magnetic resonance imaging (dMRI) or functional communication patterns derived from functional imaging modalities such as functional MRI (fMRI) or magnetoencephalography (MEG) [7, 8]. Following the construction of a brain network using one of these modalities, its topological properties can be determined through numerous metrics. Broadly speaking, these metrics fall into one of two categories: network segregation or network integration. A balance between integrative and segregative organization is considered optimal for healthy brain functioning [9, 10].

Disturbances in this optimal network organization in MS may relate to clinical impairment, and is hypothesized to center around a network collapse [11]. As structural damage accumulates, the network becomes less and less efficient until a critical point is reached, after which the network collapses and (cognitive) impairment emerges. Disruptions of network organization are well-established in MS, as evidenced by studies using dMRI, fMRI, and MEG [12, 13]. The relevance of variations in directionality and magnitude of these disturbances, however, is inconclusive within and across modalities, and as of yet there is no consensus on a common network correlate of cognitive dysfunction in MS.

These mixed results in the field may at least in part be explained by the relatively simple way previous work has looked at the brain network, i.e. by focusing on single-layer networks derived from a single modality. Such unimodal approaches disregard the crucial interplay between structure and different aspects of brain function in the brain network. For instance, we know that brain structure partly constrains functional dynamics but does not explain it fully [14, 15], suggesting a complex synergy between structure and function, which has not been evaluated extensively yet in MS. Indeed, a recent exploration looking at changes in the “core” of the network showed that incorporating information from both structure and function improved correlations with cognition [16].

The multilayer network framework provides a novel solution to study the topology of the entire brain network by integrating structure and function in one model [17–19]. Multilayer approaches have been validated previously, showing clinical relevance in schizophrenia [20, 21] as well as Alzheimer’s disease [22, 23]. In the healthy brain, we have previously used multilayer networks integrating structural MRI, functional MRI, and MEG data to show a positive relation between integration of the frontoparietal network (FPN) and executive functioning, which is less pronounced when considering isolated single-layer networks [24]. We further validated the relevance of the FPN in the multilayer in a glioma population, observing this same positive correlation with cognition when integrating the different MEG frequency bands into a multilayer network [25]. These findings suggest that such a multilayer approach may be more sensitive than unimodal analyses to individual differences in cognition across different populations.

The aim of the present work was to investigate whether the positive relation between (multilayer) integration of the FPN and cognitive functioning also holds for MS and whether multilayer network correlates supersede their single-layer counterparts.

## 2. Methods

### 2.1 Participants

For this study, we used a large cross-sectional dataset previously collected for a multimodal analysis of network dysfunction in MS [16] across 13 European centers part of the MAGNIMS consortium (http://www.magnims.eu/). The MAGNIMS study was reviewed and approved by each participating center’s local ethical committee, and written informed consent was provided by each participant independently at each center.

A total of 1517 participants were included in the study. Of these, as reported elsewhere [16], 33 were previously excluded due to suboptimal MRI quality or failures in image processing. The final dataset thus comprised 1484 participants, of which 41 people with clinically isolated syndrome, 1007 people with definite MS (according to the 2010 McDonald criteria), and 436 healthy controls (HC). Of the people with MS (pwMS), 817 had relapsing-remitting (RR) MS; 121 had secondary progressive MS; and 69 had primary progressive MS. MRI and clinical data were collected in all patients; for HC, only MRI data were obtained. Because we aimed to reduce effects of clinical heterogeneity, we focused only on the RRMS population in the present work [26, 27].

### 2.2 Neuropsychological and neurological evaluation

Clinical evaluation of pwMS was carried out around the time of MRI acquisition. To assess cognitive performance, the Symbol Digit Modalities Test (SDMT) [28] was used. The SDMT is a neuropsychological test where the participant is asked to match symbols to digits ranging from 1 through 9 according to a provided key. Here, the spoken version of this test was used to circumvent the impact of motor impairment. Although traditionally seen as a measure of information processing speed in pwMS [29], the SDMT also quantifies aspects of working memory [30, 31] and executive functioning [32]. Importantly, the SDMT has been observed to be the most frequently impacted test in MS and is a sensitive marker of overall cognitive decline in pwMS [29, 33]. Country-specific normative data were used to convert raw SDMT scores to z-scores by adjusting for gender, age, and education. To assess overall physical disability, the Expanded Disability Status Scale (EDSS) [34] was used.

### 2.3 MRI

MRI data was obtained from all participants using 3T MRI scanners, and included a 3D T1-weighted sequence, a T2-weighted fluid attenuated inversion recovery (FLAIR) sequence, and resting-state (rs) functional MRI (fMRI) and (single- and multi-shell) dMRI acquisitions. Exact acquisition protocols differed across the centers. The details of the imaging protocols, as well as the exact preprocessing of the data, are described in detail previously [16], but summarised briefly below

#### 2.3.1 Structural MRI

T2-hyperintense lesions segmented on FLAIR images were used to fill in lesions on T1w images, so that total lesion volume (TLV) could be computed. T1w images were segmented into grey matter (GM), white matter (WM), and CSF, and were used to parcellate the brain into 100 cortical Schaefer atlas regions. This atlas integrates local gradient and global similarity approaches and its parcels are assigned to seven canonical resting-state networks [35], comprised of the visual (VIS), somatomotor (SM), limbic (L), dorsal attention (DAN), ventral attention (VAN), default mode (DMN), and control or frontoparietal (FPN) networks. An additional 14 subcortical GM regions were segmented using FSL-FIRST and added to the parcellation, for a total of 114 regions.

#### 2.3.2 Resting-state functional MRI

Preprocessing of rsfMRI data was done using fMRIPrep (20.2.6, [36]), and included susceptibility induced distortion correction based on available fieldmap sequences (phase-encoding polarity method, phase-difference B0 estimation), or a registration-based fieldmapless estimation, registration to the T1w image, slice-timing correction, head motion estimation, and confound estimation. Denoising consisted of removal of non-steady state volumes, band-pass filtering (0.008-0.08Hz), detrending, standardization and confound regression (mean WM and CSF signal and non-aggressive ICA-AROMA components).

Time series were extracted from all atlas regions and Pearson correlation coefficients between all pairs of time series were computed, Fisher z-transformed, and absolutized to obtain a 114 by 114 functional connectivity matrix.

#### 2.3.3 Diffusion MRI

Preprocessing of dMRI was performed using QSIPrep (0.14.3, [37]), and included denoising using MP-PCA, distortion correction using available sequences, head motion and eddy current correction, and resampling to the T1w image. A tissue response function was estimated using the Dhollander algorithm, and the fiber orientation distribution (FOD) for each voxel was determined using constrained spherical deconvolution and subsequently intensity-normalized using mtnormalize. Probabilistic anatomically-constrained tractography based on WM FODs was used to generate 10 million streamlines and spherical-deconvolution informed filtering of tractograms (SIFT2, [38]) was then performed to obtain weights for each streamline. A 114 by 114 structural connectivity matrix was obtained by summing the weights of all streamlines between each pair of atlas regions. To make the distribution of structural connectivity link weights more comparable to functional connectivity link weights, structural connectivity matrices were log10-transformed.

#### 2.3.4 Cross-site harmonization

As data were acquired using different MRI systems and acquisition protocols in 13 centers across Europe, ComBat harmonization [39] was used to correct structural and functional connectivity matrices and brain volumes for center-specific effects while maintaining biological associations with gender, age, and education.

### 2.4 Network construction

#### 2.4.1 Single-layer networks

First, we used MATLAB (R2022b, Mathworks, Natick, MA, USA) to construct minimum spanning trees (MST) for the structural and functional single-layer networks by applying Kruskal’s algorithm [40]. Briefly, links were ordered by weight (strongest to weakest) after which all the nodes in the network were connected using only the strongest links without forming any loops, yielding the strongest subgraph of the original graph (assuming network weights are distinct). Remaining links were then binarized. The use of the MST prevents common issues stemming from differences in link density or connection strength across layers and between subjects [41, 42]. We used functions of the brain connectivity toolbox (2019.03.03, https://sites.google.com/site/bctnet/) to compute several single-layer network metrics for the functional and structural networks (see Methods 2.5).

#### 2.4.2 Multilayer networks

We then integrated the structural and functional matrices in a multimodal multiplex, a multilayer network in which links between layers were present only between a node’s counterpart across layers. In keeping with the intralayer links (i.e. the links *within* a single-layer network of the multiplex), these interlayer links (i.e. the links *between* the single-layer networks of the multiplex) were binarized. A multiplex thus comprised L = 2 layers (one for rsfMRI and one for dMRI) where each layer consisted of an identical set of N = 114 nodes (the atlas regions) and M = N – 1 = 113 links. Multiplexes were represented as LxN by LxN supra-adjacency matrices where the diagonal NxN blocks encode intralayer connectivity and off-diagonal NxN blocks encode interlayer connectivity. These supra-adjacency matrices were exported to Python (3.6, Python Software Foundation, available at http://python.org/) where we used in-house scripts that implement the Python libraries NetworkX (2.3, https://github.com/networkx) and multiNetX (https://github.com/nkoub/multinetx) to compute several multilayer network metrics. All scripts used to analyze the data are openly available on our GitHub page at https://github.com/multinetlab-amsterdam. Figure 1 shows a schematic overview of the analysis pipeline.

**Figure 1.**
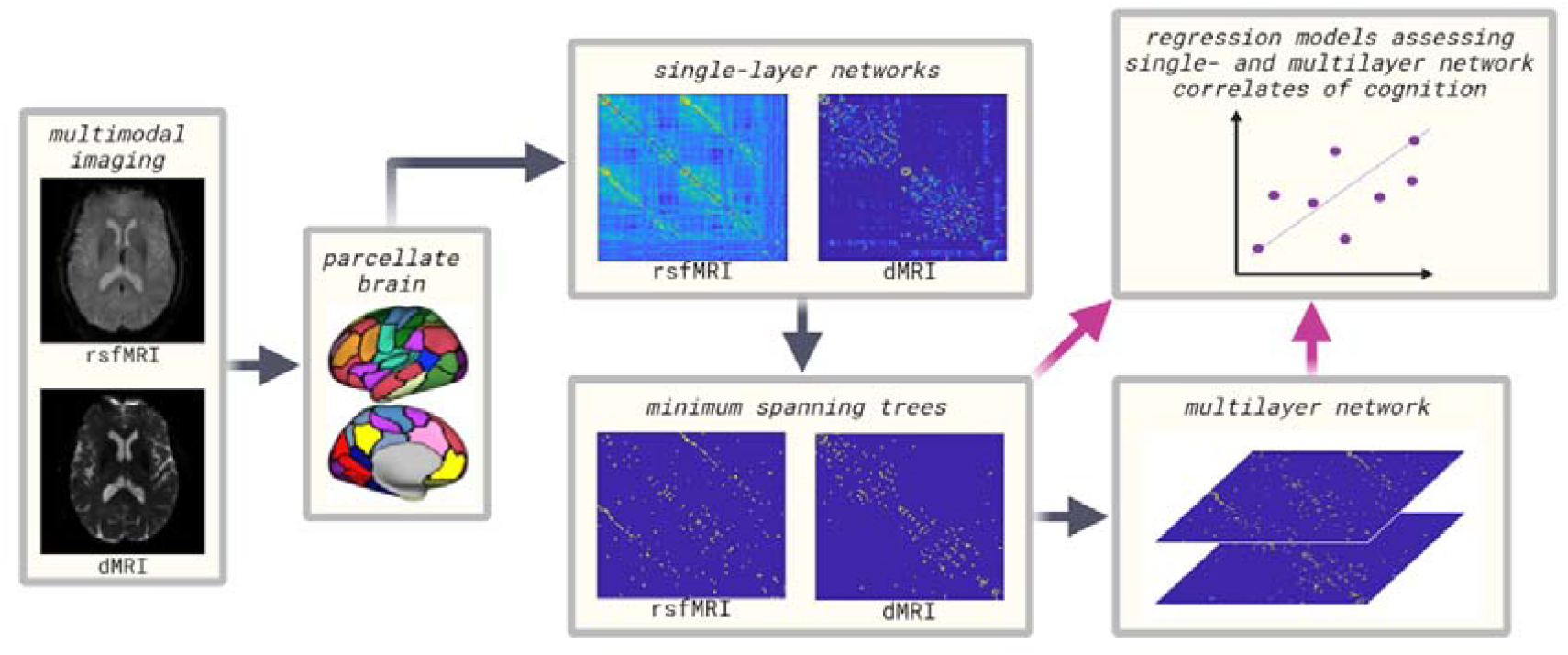
Flowchart of the analysis pipeline.

### 2.5 Network metrics

#### 2.5.1 Main analysis

Eigenvector centrality (*EC*) is a centrality metric that takes into account not only the connections of a node itself but also that of its neighbors [43]. We computed nodal *EC* for the structural and functional single-layer networks and the multiplexes. For all three networks separately, *EC* values of nodes belonging to the FPN were extracted and averaged to obtain one value per subject representing *EC* of the FPN (*ECfpn*).

#### 2.5.2 Post-hoc

To further elucidate the single- and multilayer network correlates of cognition in pwMS, we also explored the mean network eccentricity. Eccentricity is a nodal measure obtained by computing the longest shortest path between a specific node and any other node in a network [44]; mean eccentricity is a global measure reflecting the average distance information has to travel through a network. We computed mean eccentricity of the entire network for the structural and functional single-layer networks as well as the multiplex.

### 2.6 Statistical analyses

#### 2.6.1 Main analysis

We tested our primary hypothesis on the relation between *ECfpn* and cognition (SDMT z-score) through a hierarchical multiple regression. Covariates (age and gender) were entered in block 1, single-layer functional and structural *ECfpn* were entered in block 2, and multilayer *ECfpn* was entered in block 3. Multicollinearity was checked for through observation of bivariate correlations between all independent variables as well as running collinearity diagnostics. All statistical analyses were performed in SPSS (version 28.0.1.1, IBM Corp., Armonk, NY, USA) using a two-tailed significance threshold of *p* < 0.05.

#### 2.6.2 Post-hoc analyses

To assess the association between mean eccentricity and the SDMT, an additional hierarchical multiple regression was run.

Furthermore, we previously showed an inverted-U relationship between age and multiplex centrality of the FPN in a healthy population when utilizing a multilayer network comprised of dMRI, rsfMRI, and MEG [24], indicating a non-linear relation between multilayer parameters and age. To validate these findings in the current large HC population, we probed the biological relevance of *ECfpn* in a structure-function multiplex by running another hierarchical multiple regression. In this model, *ECfpn* was the dependent variable and age and the square of age were entered in a first and second block, respectively.

Next, to ensure the robustness of our findings, we repeated all analyses using an alternative 224-region brain parcellation: the 210 cortical regions of the Brainnetome atlas (BNA) [45] combined with the 14 FSL-FIRST-derived subcortical GM regions. We used an earlier categorization [46] of the BNA parcels into the seven canonical resting-state networks to determine FPN nodes for the computation of *ECfpn*.

We performed a final additional validation step using another dataset, namely the MuMoBrain dataset consisting of 33 HC where we previously observed an association between multilayer FPN integration and cognition as well as age [24]. See the Supplementary Materials for a brief description of this dataset. For these 33 HC, we obtained structure-function (dMRI/fMRI) multiplex networks constructed in identical manner as described in the current work, and used regression models to again relate FPN integration of these multilayers to cognition and age.

## 3. Results

### 3.1 Participants

After exclusion of participants with missing data on any relevant variables, i.e. cognition, age, gender, or EDSS, a total of 780 people with RRMS remained. Information on age and gender was available for all 436 HC. Characteristics of both samples are described in Table 1. Of all pwMS, 200 (25.6%) showed impairment of the SDMT at a z-score < −1.5. The median EDSS was 2.0 (IQR 1.0-3.0).

**Table 1.**
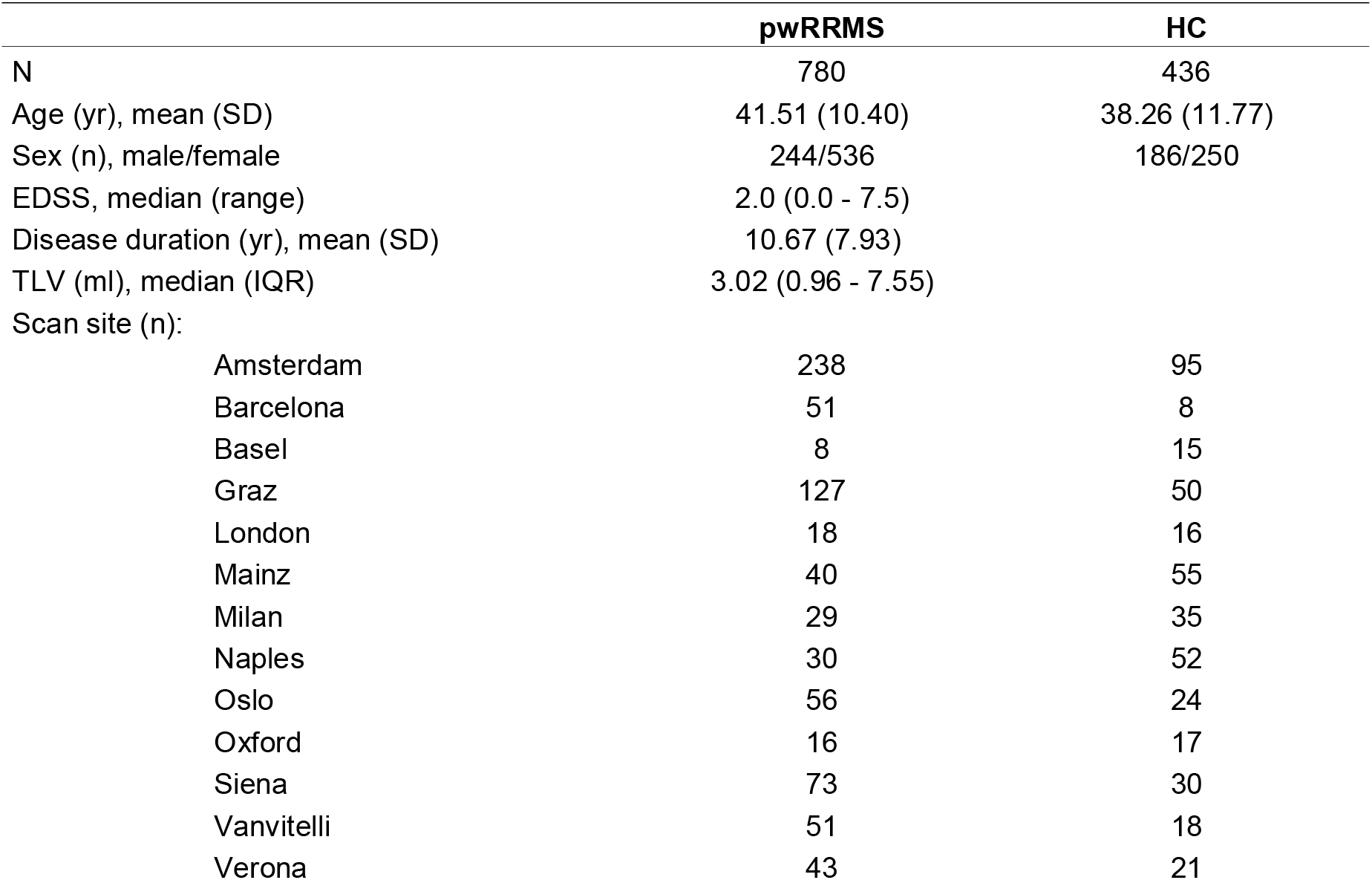
Demographics of the patients with RRMS as well as the healthy controls. pwRRMS = people with relapsing-remitting multiple sclerosis. HC = healthy controls. EDSS = expanded disability status scale. TLV = total lesion volume.

|  | pwRRMS | HC |
| --- | --- | --- |
| N | 780 | 436 |
| Age (yr), mean (SD) | 41.51 (10.40) | 38.26 (11.77) |
| Sex (n), male/female | 244/536 | 186/250 |
| EDSS, median (range) | 2.0 (0.0 - 7.5) |  |
| Disease duration (yr), mean (SD) | 10.67 (7.93) |  |
| TLV (ml), median (IQR) | 3.02 (0.96 - 7.55) |  |
| Scan site (n): |  |  |
| Amsterdam | 238 | 95 |
| Barcelona | 51 | 8 |
| Basel | 8 | 15 |
| Graz | 127 | 50 |
| London | 18 | 16 |
| Mainz | 40 | 55 |
| Milan | 29 | 35 |
| Naples | 30 | 52 |
| Oslo | 56 | 24 |
| Oxford | 16 | 17 |
| Siena | 73 | 30 |
| Vanvitelli | 51 | 18 |
| Verona | 43 | 21 |

### 3.2 Cognition and eigenvector centrality of the frontoparietal network: The added value of multiplex measures

Our data showed that gender, age, and multiplex *ECfpn* were significantly associated with cognitive performance, and that the full model of gender, age, single-layer structural and functional *ECfpn*, and multiplex *ECfpn* was statistically significant (*R*^*2*^ = .054, adjusted *R*^*2*^ = .048, *F*[5, 774] = 8.880, *p* < .001). However, single-layer structural and functional *ECfpn* did not significantly add to the model, and the model only explained 4.8% of variance in SDMT scores. Moreover, the level of integration of the FPN in the multiplex was not positively but negatively related to SDMT scores in pwMS (β = −.117, *p* = .005). See Table 2 and Figure 2.

**Table 2.** Hierarchical multiple regression predicting cognitive performance from single- and multilayer EC of the frontoparietal network** * p<.05, ** p<.001.

| Variable | Model 1 |  | Model 2 |  | Model 3 |  |
| --- | --- | --- | --- | --- | --- | --- |
| | B | $\beta$ | B | $\beta$ | B | $\beta$ |
| (Constant) | 0.419* |  | 0.417* |  | 0.560* |  |
| Age | -0.023** | -0.179 | -0.023** | -0.180 | -0.023** | -0.180 |
| Sex | -0.323* | -0.112 | -0.319* | -0.111 | -0.332* | -0.116 |
| $EC_{fpn}$ single-layer structural network | | | -0.330 | -0.012 | 0.510 | 0.018 |
| $EC_{fpn}$ single-layer functional network | | | 0.365 | 0.021 | 1.359 | 0.079 |
| $EC_{fpn}$ multiplex | | | | | -1.552* | -0.117 |
| $R^2$ | 0.044 | | 0.045 | | 0.054 | |
| $F$ | 17.959** | | 9.080** | | 8.880** | |
| $\Delta R^2$ | 0.044 | | 0.001 | | 0.009 | |
| $\Delta F$ | 17.959** | | 0.236 | | 7.764* | |

\* $p < .05$ , \*\* $p < .001$

**Figure 2.**
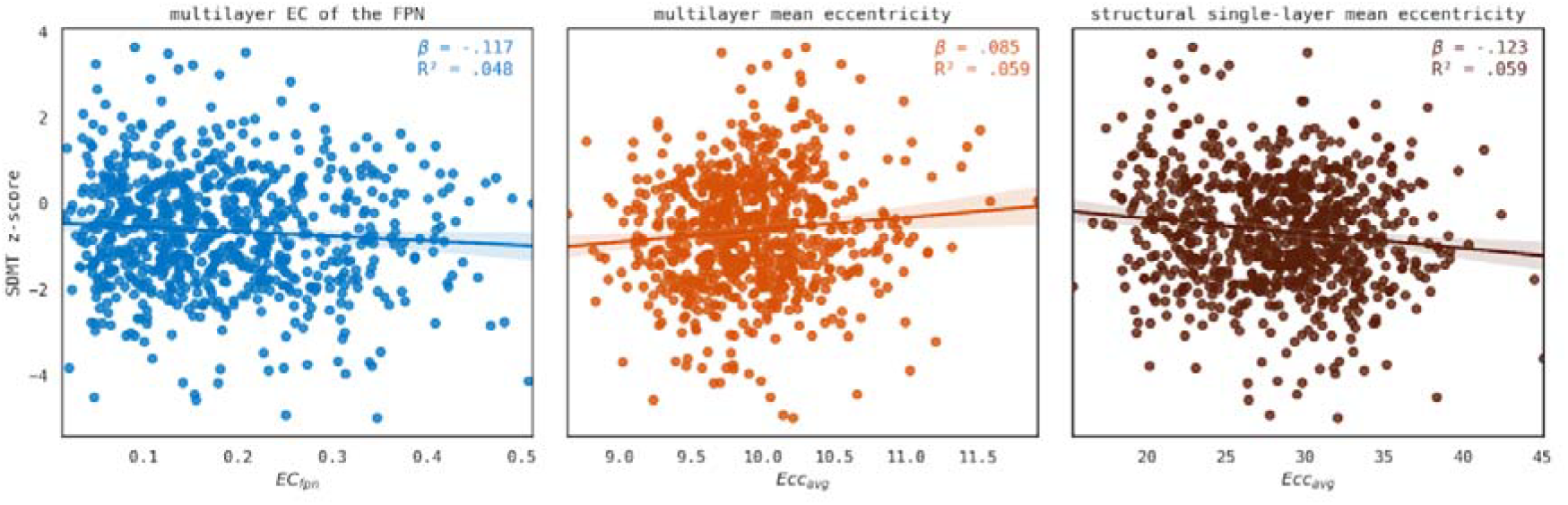
Network measures and cognition in MS. Scatterplots including lines of best fit of significant associations between network correlates and SDMT performance. From left to right: multilayer eigenvector centrality of the frontoparietal network; mean eccentricity of the multilayer network; mean eccentricity of the single-layer structural network. SDMT = symbol-digit modalities test. EC = eigenvector centrality. FPN = frontoparietal network. EC_fpn_ = eigenvector centrality of the frontoparietal network. Ecc_avg_ = mean eccentricity.

### 3.3 Post-hoc evaluations: Network eccentricity, age, and validation analyses

#### 3.3.1 Cognition and mean eccentricity in MS

Mean eccentricity was associated with SDMT scores (*R*^*2*^ = .065, adjusted *R*^*2*^ = .059, *F*[5, 774] = 10.804, *p* < .001; see Supplementary Table 1 and Figure 2). A shorter average longest path length in the single-layer structural network (β = −.123, *p* < .001) but longer average path length in the multilayer network (β =0.085, *p* = .018) related to better SDMT scores in pwMS. However, effects were small, explaining only 5.9% of the variance in SDMT scores.

#### 3.3.2 Age and multilayer eigenvector centrality of the frontoparietal network in HC

In contrast to our previously observed non-linear relation between age and multilayer parameters in HC (see Methods 2.6.2), a regression model in the current HC population revealed that multilayer *ECfpn* in a structure-function multiplex, in fact, did not follow the expected quadratic relationship with age (*p* = .788; see Supplementary Table 1 and Figure 2).

#### 3.3.3 Validation analyses: Brainnetome atlas

Although the full models assessing the relation between SDMT scores and *ECfpn* and mean eccentricity were significant (*p* < .001), in both models age and gender were the only significant predictors of executive functioning, see Supplementary Table 2. The single- and multilayer network metrics added no significant value to the models.

For the association between age and multilayer *ECfpn*, results of the full model were non-significant (*p* = .217) and thus similar to the analysis using the Schaefer atlas. Neither age (β = −.484, *p* = .177) nor age squared (β = .427, *p* = .235) were significant predictors of multilayer *ECfpn*, indicating no relevant relation between a structure-function network correlate and age.

#### 3.3.4 Validation analyses: MuMoBrain dataset

In summary, our results showed a weak correlation between multilayer measures and cognition that could not be replicated when using a different atlas. Interestingly, FPN integration of the MuMoBrain structure-function multilayer without MEG similarly showed no significant relation with cognition (*R*^*2*^ = .065, adjusted *R*^*2*^ = −.032, *F*[3, 29] = .673, *p* = .576) nor age (*R*^*2*^ = .031, adjusted *R*^*2*^ = −.034, *F*[2, 30] = .478, *p* = .625), see Supplementary Table 3 and Supplementary Figure 1; Supplementary Figure 2 shows the significant associations present in the original multilayer with MEG.

## 4. Discussion max 1600 words (now 1267)

In this study, we aimed to investigate whether multilayer frontoparietal network parameters, integrating structural and functional MRI data, were related to cognition in MS. This aim was chosen based on earlier observations of a positive relation with cognition in both HCs [24] and glioma patients [25].

However, in this dataset, we found a weak and negative relation between integration of the FPN in the multilayer network and cognition in pwMS. These findings could not be replicated when using a different atlas. Additional validation analyses suggest that our findings may be affected by the absence of MEG in this dataset, in contrast to previous studies [24, 25].

We did not observe a consistent relation between multilayer parameters and cognition in pwMS. Interestingly, a recent study in a largely overlapping dataset did show a relation with cognition for other measures, looking at hub-specific patterns of integration between structure and function; but this analysis used a less synergistic approach [16]. That study used an additive multimodal approach, not directly considering the links between the structural and functional layers. Furthermore, the study also included an additional structural layer in the form of morphological covariance networks. The analysis could therefore have been driven by a predominance of structural layers. Moreover, morphological networks are based on the covariance of grey matter volumes between brain regions, and may thus also reflect the grey matter damage, which is a main hallmark of MS and relates to clinical outcomes [47, 48]. This was not included in our study, as our approach was based on the aforementioned HC and glioma studies, where morphological networks were also not present. Another reason for the absence of a significant relation with cognition could reside in specific methodological choices we made. For instance, interlayer links were present only between identical node pairs across layers and were binarized (i.e. with a weight of 1) across the entire network. This approach fails to acknowledge the heterogeneity of (dMRI-fMRI) structure-function tethering across the cortex, which, moreover, shows low convergence in higher-order areas such as the FPN [49]. In neurodegenerative populations including pwMS, this structure-function coupling may be even more complex and more difficult to capture in a single measure [50, 51]. When integrating these modalities, it may therefore be crucial to take into account the coupling between structure and function, particularly in those parts of the brain where this relationship is inherently weak.

We speculate that the negative relation we see between multilayer integration of the FPN and SDMT scores is a sign of a pathological network organization in MS. While strong FPN integration into the entire brain network is integral for cognition and executive functions in particular [24, 52–54], it is just as necessary that highly central hub regions, such as those in the FPN, are counterbalanced by less central non-hub regions [9, 10]. Without this equilibrium, the load on the central nodes becomes excessive, leading to network overload and subsequent failure and concomitant cognitive impairment [55]. Indeed, previous studies in MS have shown heightened centrality of hubs in cognitively impaired patients, including in the FPN [56–58]. This is further supported by our analyses of network eccentricity, showing that higher average path length in the multilayer relates to better cognition. If we assume central node overload leads to disturbed cognition in these pwMS, short path lengths in the MSTs of pwMS with worse cognitive performance might similarly indicate a starlike network organization that leans too heavily on a few central nodes, rendering the network vulnerable to overload and failure. In contrast, the much higher eccentricity in the single-layer diffusion network may indicate a network configuration that, potentially due to structural damage caused by demyelination, has become too line-like—lacking the central hub nodes needed for efficient information processing— and thus similarly inefficient. However, as effect sizes of associations between network measures and cognitive performance were small and validation analyses were non-significant, future research replicating the relations between multilayer integration of the FPN and SDMT scores in pwMS is necessary.

We consistently find significant associations between cognitive performance and gender and age: men performed slightly worse than women, and older age related to worse cognition. While these findings might seem counterintuitive initially given that the SDMT scores have been corrected for demographic variables, normscoring corrects only for the normal variation in these scores that can be attributed to age and gender. It does not, however, correct for disease-specific variations. Indeed, men suffering from MS commonly present with more cognitive dysfunction than women [59–61]. Similarly, longer disease duration, which correlates with, and is difficult to disentangle from age, is linked to worse cognitive performance [27, 60, 62].

Our study also aimed to validate previous findings in HC, namely the previously observed non-linear relation between multilayer FPN integration and age [24]. A U-shaped relation with age is ubiquitous in many other brain measures, such as whole brain- [63] and white matter volumes [64, 65], but also network efficiency [66–68], all of which increase during early life and subsequently decline with older age. We did not observe such a relation with age in HC in the present MAGNIMS dataset. However, the previous HC dataset in which we did observe this relation included more functional modalities, i.e. MEG, in the multilayer network [24]. Interestingly, this relation with age was no longer present after excluding MEG. We may speculate that this reflects the importance of different functional data in the multilayer context, and particularly for cognition. The specific relevance of MEG beyond fMRI could be explained by the fact that MEG can capture properties of faster neuronal oscillations which may be particularly sensitive to cognitive functions [69]. Another consideration is the dominance of functional layers in the MuMoBrain study versus the present work: our current multilayer was comprised of 50% functional data, while the significant correlations between cognition and integration in both healthy controls and glioma patients were based on multilayers that were comprised of 88-100% functional data. The fact that reanalysis of the MuMoBrain HC data without the additional functional MEG layers yielded no significant correlations with age and cognition further supports the potential relevance of incorporating additional functional modalities in multimodal explorations of cognition in MS, and opens up interesting avenues for future research.

Several limitations of this work should be acknowledged. In our previous work, executive functioning was the main outcome of interest, and was quantified through multiple neuropsychological tests. In the present MS dataset, however, only a single assessment of more general cognitive performance (i.e. the SDMT) was available. Additionally, given the multi-center nature of this dataset, inter-site differences in test administration and normative populations used for z-scoring of SDMT test performance should be noted, as should differences in specific MRI scanners and acquisition protocols; although we did use statistical harmonization techniques to minimize effects of the latter. Furthermore, the binarized multiplex we employed does not take into account link weights or interactions between different regions across modalities; future work may look to explore alternative methods. Finally, in order to reduce clinical heterogeneity, the present study investigated only people with RRMS, while specific evaluations of changing relations between network measures and cognition across the disease span could be of subsequent interest.

To conclude, we observed network correlates of cognition using a structure-function multilayer network approach in MS. However, observed effect sizes were negligible and results did not survive validation analyses, potentially highlighting the shortcomings of a binary structure-function multilayer comprised of only dMRI and fMRI. Our study underscores the importance of further exploring the complex interplay between multimodal neuroimaging network dynamics and cognition, and future studies may consider additional (functional) modalities or alternative multilayer approaches to further elucidate these associations.

## Data Availability

Data from patients are controlled by the respective centers (listed in Supplementary Table 1) and therefore are not publicly available. Request to access raw data should be forwarded to data controllers via the corresponding author. Derived data supporting the findings of this study can be requested by qualified investigators from the corresponding author.

## Conflicts of Interest

**FB:** Steering committee and iDMC member for Biogen, Merck, Roche, EISAI. Consultant for Roche, Biogen, Merck, IXICO, Jansen, Combinostics. Research agreements with Novartis, Merck, Biogen, GE, Roche. Co-founder and share-holder of Queen Square Analytics LTD.

**AB** has received speaker honoraria and/or compensation for travel grant and consulting service from Biogen, Merck, Genzyme, Novartis, Alexion, Amgen, UCB, Coloplast and Roche.

**AC** is supported by EUROSTAR E!113682 HORIZON2020 and received speaker honoraria from Novartis and Roche.

**MC** received speaker honoraria from Biogen, Bristol Myers Squibb, Celgene, Genzyme, Merck Serono, Novartis, and Roche and receives research support from the Progressive MS Alliance and Italian Minister of Health.

**OC** is an NIHR Research Professor (RP-2017-08-ST2-004); acts as a consultant for Biogen, Merck, Novartis, Roche, and Teva; and has received research grant support from the MS Society of Great Britain and Northern Ireland, the NIHR UCLH Biomedical Research Centre, the Rosetree Trust, the National MS Society, and the NIHR-HTA.

**SC** received travel support and speaker honoraria from Merck and is supported by Rosetrees Trust (MS632; PGL21/10079)

**RC** was awarded a MAGNIMS-ECTRIMS fellowship in 2019; she received speaker honoraria/travel support from Roche, Merck Serono, UCB, Sanofi-Genzyme, Alexion, Novartis, and Janssen, and received a research grant from the Italian Ministry of University and Research.

**CE** received travel funding and speaker honoraria from Biogen Idec, Bayer Schering Pharma, Merck Serono, Novartis, Genzyme and Teva Pharmaceutical Industries Ltd./Sanofi-Aventis, Shire; received research support from Merck Serono, Biogen Idec, and Teva Pharmaceutical Industries Ltd./Sanofi-Aventis; and serves on scientific advisory boards for Bayer Schering Pharma, Biogen Idec, Merck Serono, Novartis, Genzyme, Roche, and Teva Pharmaceutical Industries Ltd./Sanofi-Aventis.

**MF** is Editor-in-Chief of the Journal of Neurology, Associate Editor of Human Brain Mapping, Neurological Sciences, and Radiology; received compensation for consulting services from Alexion, Almirall, Biogen, Merck, Novartis, Roche, Sanofi; speaking activities from Bayer, Biogen, Celgene, Chiesi Italia SpA, Eli Lilly, Genzyme, Janssen, Merck-Serono, Neopharmed Gentili, Novartis, Novo Nordisk, Roche, Sanofi, Takeda, and TEVA; participation in Advisory Boards for Alexion, Biogen, Bristol-Myers Squibb, Merck, Novartis, Roche, Sanofi, Sanofi-Aventis, Sanofi-Genzyme, Takeda; scientific direction of educational events for Biogen, Merck, Roche, Celgene, Bristol-Myers Squibb, Lilly, Novartis, Sanofi-Genzyme; he receives research support from Biogen Idec, Merck-Serono, Novartis, Roche, the Italian Ministry of Health, the Italian Ministry of University and Research, and Fondazione Italiana Sclerosi Multipla

**MAF** has received speaker honoraria from Merck.

**CG**: The University Hospital Basel (USB) and the Research Center for Clinical neuroimmunology and Neuroscience (RC2NB), as the employers of Cristina Granziera, have received the following fees which were used exclusively for ( research support from Siemens, GeNeuro, Genzyme-Sanofi, Biogen, Roche. They also have received advisory board and consultancy fees from Actelion, Genzyme-Sanofi, Novartis, GeNeuro, Merck, Biogen and Roche; as well as speaker fees from Genzyme-Sanofi, Novartis, GeNeuro, Merck, biogen and Roche.

**EAH** received honoraria for advisory board activity from Sanofi-Genzyme, and his department has received honoraria for lecturing from Biogen and Merck.

**SL** received speaker honoraria from Sanofi, Biogen, Bristol Myers Squibb, Novartis and Merck.

**SM** received speaking honoraria from UCB, and travel grants from Sanofi, Merck, Alexion, UCB and Roche.

**MM** received research funding from MUR PNRR Extended Partnership (MNESYS no. PE00000006, and DHEAL-COM no. PNC-E3-2022-23683267), ECTRIMS-MAGNIMS, UK MS Society, e Merck; salary as editorial board member from Neurology (AAN, MN, USA), and Multiple Sclerosis Journal (Sage, UK); and honoraria from Abbvie, Biogen, BMS Celgene, Ipsen, Jansen, Merck, Novartis, Roche, and Sanofi-Genzyme.

**JP** has received support for scientific meetings and honorariums for advisory work From Merck Serono, Novartis, Chugai, Alexion, Roche, Medimmune, Argenx, Vitaccess, UCB, Mitsubishi, Amplo, Janssen. Grants from Alexion, Argenx, Roche, Medimmune, Amplo biotechnology. Patent ref P37347WO and licence agreement Numares multimarker MS diagnostics Shares in AstraZenica. Her group has been awarded an ECTRIMS fellowship and a Sumaira Foundation grant to start later this year. A Charcot fellow worked in Oxford 2019-2021. She acknowledges partial funding to the trust by Highly specialised services NHS England. She is on the medical advisory boards of the Sumaira Foundation and MOG project charities, is a member of the Guthy Jackon Foundation Charity and is on the Board of the European Charcot Foundation and the steering committee of MAGNIMS and the UK NHSE IVIG Committee and chairman of the NHSE neuroimmunology patient pathway and ECTRIMS Council member on the educational committee since June 2023. On the ABN advisory groups for MS and neuroinflammation.

**DP** is a member of the advisory board for “Cognition and MS” for Novartis and received speaking honoraria from Biogen, Novartis, MedAhead and Bristol-Myers Squibb.

**GP** received research grants from ECTRIMS, MAGNIMS, and ESNR

**MAR** received consulting fees from Biogen, Bristol Myers Squibb, Eli Lilly, Janssen, Roche; and speaker honoraria from AstraZaneca, Biogen, Bristol Myers Squibb, Bromatech, Celgene, Genzyme, Horizon Therapeutics Italy, Merck Serono SpA, Novartis, Roche, Sanofi and Teva. She receives research support from the MS Society of Canada, the Italian Ministry of Health, the Italian Ministry of University and Research, and Fondazione Italiana Sclerosi Multipla. She is Associate Editor for Multiple Sclerosis and Related Disorders; and Associate Co-Editor for Europe and Africa for Multiple Sclerosis Journal

**MMS** serves on the editorial board of Neurology and Frontiers in Neurology, receives research support from the Dutch MS Research Foundation, Eurostars-EUREKA, ARSEP, Amsterdam Neuroscience, MAGNIMS and ZonMW and has served as a consultant for or received research support from Atara Biotherapeutics, Biogen, Celgene/Bristol Meyers Squibb, Genzyme, MedDay and Merck.

**AT** has received speaker honoraria from Merck, Biomedia, Sereno Symposia International Foundation, Bayer and At the Limits and meeting expenses from Merck, Biogen Idec and Novartis. He is co-editor for Multiple Sclerosis Journal, an associate editor for Frontiers in Neurology – Neuro-ophthalmology section and on the editorial board for Neurology. He has been supported by recent grants from the MRC (MR/S026088/1), NIHR BRC (541/CAP/OC/818837) and RoseTrees Trust (A1332 and PGL21/10079).

**PV** received speaker honoraria from Biogen Idec.

The remaining authors report no competing interests.

## Notes

### Funding Statement

G.P. was supported by the ECTRIMS-MAGNIMS Research Fellowship Programme (2020). F.P. and B.K. are supported by the UK National Institute for Health Research (NIHR) Biomedical Research Centre (BRC) at UCLH and UCL. A.C. is supported by EUROSTAR E!113682 HORIZON2020. Sa.Co. is supported by a Rosetrees Trust Grant (PGL21/10079). M.A.F. is supported by a grant from the MRC (MR/S026088/1). L.D. Is supported by the Dutch Research Council (NWO, Vidi 198.015). The study was supported by grants from The Research Council of Norway (NFR, grant number 240102) and the South-Eastern Health Authorities of Norway (grant number 257955).

### Author Declarations

Ethics Committee of University College London gave approval for this work.

