## Supplemental Materials for "Structure-function multilayer network integration and cognition in multiple sclerosis"

**The MuMoBrain dataset**

To assess the relevance of a binary structure-function multiplex network, we performed a *post-hoc* analysis in the previously obtained MuMoBrain dataset [1], testing the relationship between multilayer centrality of the FPN and executive functioning (EF) and age.

***Participants***

Thirty-nine healthy volunteers were recruited for this study through Hersenonderzoek.nl (www.hersenonderzoek.nl). Participants met the following criteria: aged 20-70, native Dutch speakers, and able to give written consent. Exclusion criteria included a history of neurological or psychiatric conditions, regular use of centrally acting drugs, and contraindications for MRI or MEG. Participants were instructed to avoid caffeine and alcohol on test days. The study received approval from the VU University Medical Center Ethics Committee, and all participants gave written consent.

Out of the 39 participants, two withdrew before completing the study, two were excluded due to MRI contraindications, and two more were excluded after their MRI scans showed artifacts. This left 33 participants with full structural MRI, dMRI, rsfMRI, and neuropsychological data for analysis. The group consisted of 18 women and 15 men, with ages ranging from 22 to 70 years (mean age 46 ± 17 years).

***Neuropsychological evaluation***

Participants underwent a comprehensive neuropsychological test battery. We used subscores from three of the included tests to quantify EF. The first was the Concept Shifting Test (CST, [2]), where participants had to cross out circles in ascending, alphabetical, or alternating order depending on whether the circles contained digits (CST-A), letters (CST-B), or both (CST-C). To adjust for motor speed, a control condition with empty circles was performed three consecutive times (CST-0). The second test was the Stroop Color-Word Test (SCWT, [3]), where participants read through four cards: first, color names printed in black ink; second, colored rectangles; third, color names printed in mismatched ink, requiring participants to name the ink color rather than the word; and fourth, the same as the third, but with some words circled, where the circled words had to be read instead of the ink color. The third and final test was the Categorical Word Fluency Test [4], where participants named as many animals as possible in 60 seconds.

Raw scores from the three tests were converted into z-scores using established norms. CST scores were adjusted for age, SCWT scores were adjusted for age, education, and age squared, and Word Fluency scores were corrected for age and education. EF was calculated by taking the average of z-scores for Word Fluency, Stroop-interference (time for card 3 adjusted for time for card 2), and CST-shift (time for card C minus the average time for cards A and B, adjusted for the average time for the control condition).

***MRI***

MRI data were obtained from all participants using a 3T Philips Ingenia CX system with a 32-channel head coil. The protocol included a high-resolution 3D T1-weighted sequence, a multiband rsfMRI sequence, and a multi-shell dMRI sequence. The exact acquisition protocol, as well as the preprocessing of the data, is described in detail elsewhere [1].

Briefly, preprocessing of rsfMRI data was performed using FSL 5 (FMRIB 2012, Oxford, United Kingdom, <http://www.fmrib.ox.ac.uk/fsl>). Steps included brain extraction, discarding the first four volumes, motion correction using six motion parameters, and spatial smoothing at 5 mm FWHM. ICA-AROMA [5] was applied to remove additional motion artifacts. Signals from white matter and cerebrospinal fluid were regressed out and a high-pass filter (100 s cutoff) was applied. Timeseries were extracted from all cortical regions of the Brainnetome (BNA, [6]) atlas. Thirteen regions with signal loss due to magnetic field inhomogeneities were excluded, leaving 197 BNA regions for analysis. Pearson correlation coefficients were computed between all pairs of time series and absolutized, resulting in a 197 by 197 functional connectivity matrix.

For dMRI, we used MRtrix3 [7] to preprocess the data. A tissue response function was estimated from pre-processed, bias-corrected dMRI data using the multi-shell multi-tissue five-tissue-type algorithm (msmt_5tt), and the fiber orientation distribution (FOD) for each voxel was determined using multi-shell multi-tissue constrained spherical deconvolution [8]. Probabilistic anatomically-constrained tractography [9] was used to generate 100 million streamlines and spherical-deconvolution informed filtering of tractograms (SIFT2, [10]) was performed to improve accuracy of the streamlines and reduce false positives. We obtained a 197 by 197 structural connectivity matrix by summing the weights of all streamlines between all pairs of BNA atlas regions.

*Supplementary Figure 1.* *Structure-function multilayer (dMRI & rsfMRI) FPN integration and cognition and age in MuMoBrain.* Scatterplots including lines of best fit of (non-significant) associations between multilayer frontoparietal network centrality and executive functioning and age in the MuMoBrain dataset. Left: multilayer eigenvector centrality of the frontoparietal network and executive functioning. Right: age and multilayer eigenvector centrality of the frontoparietal network. EC = eigenvector centrality. FPN = frontoparietal network. EF = executive functioning. EC_fpn_ = eigenvector centrality of the frontoparietal network.

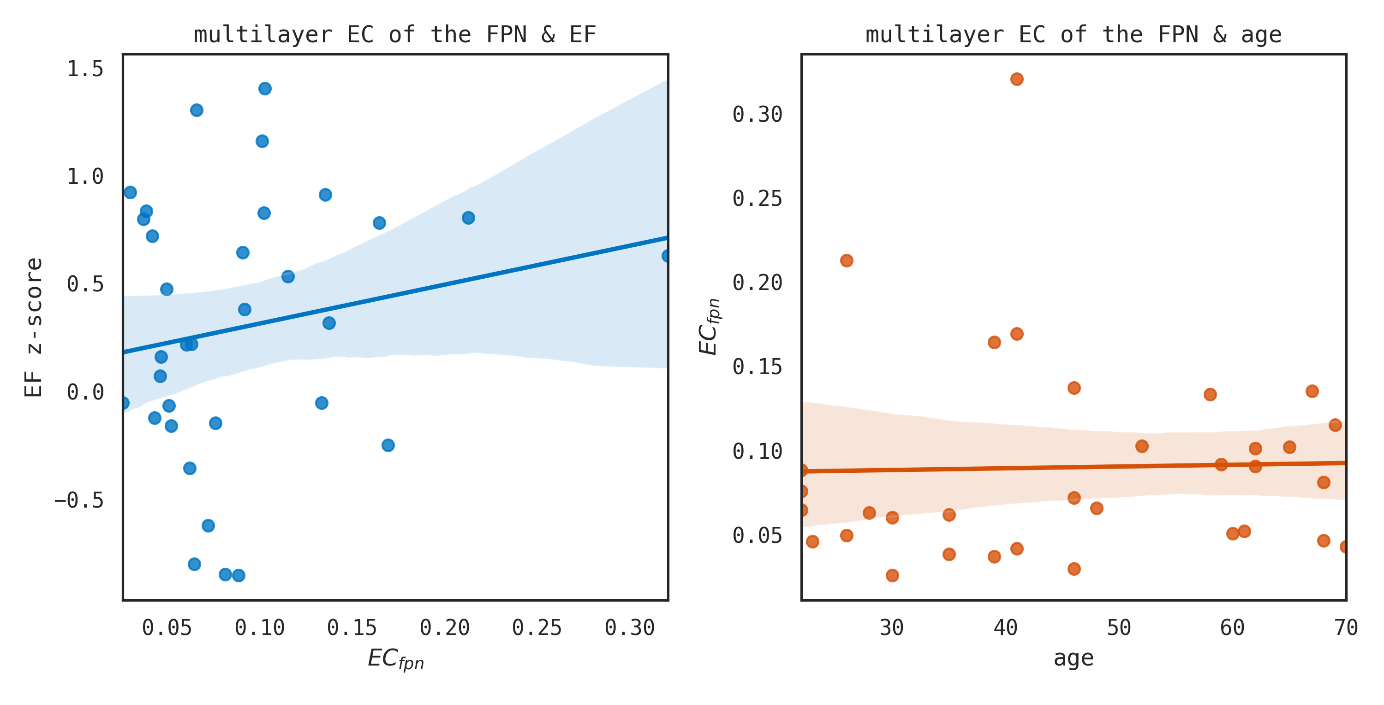

*Supplementary Figure 2.* *Multilayer (dMRI, rsfMRI, and MEG) FPN integration and cognition and age in MuMoBrain.* Scatterplots including lines of best fit of (non-significant) associations between multilayer frontoparietal network centrality and executive functioning and age in the MuMoBrain dataset. Left: multilayer eigenvector centrality of the frontoparietal network and executive functioning. Right: age and multilayer eigenvector centrality of the frontoparietal network. EC = eigenvector centrality. FPN = frontoparietal network. EF = executive functioning. EC_fpn_ = eigenvector centrality of the frontoparietal network.

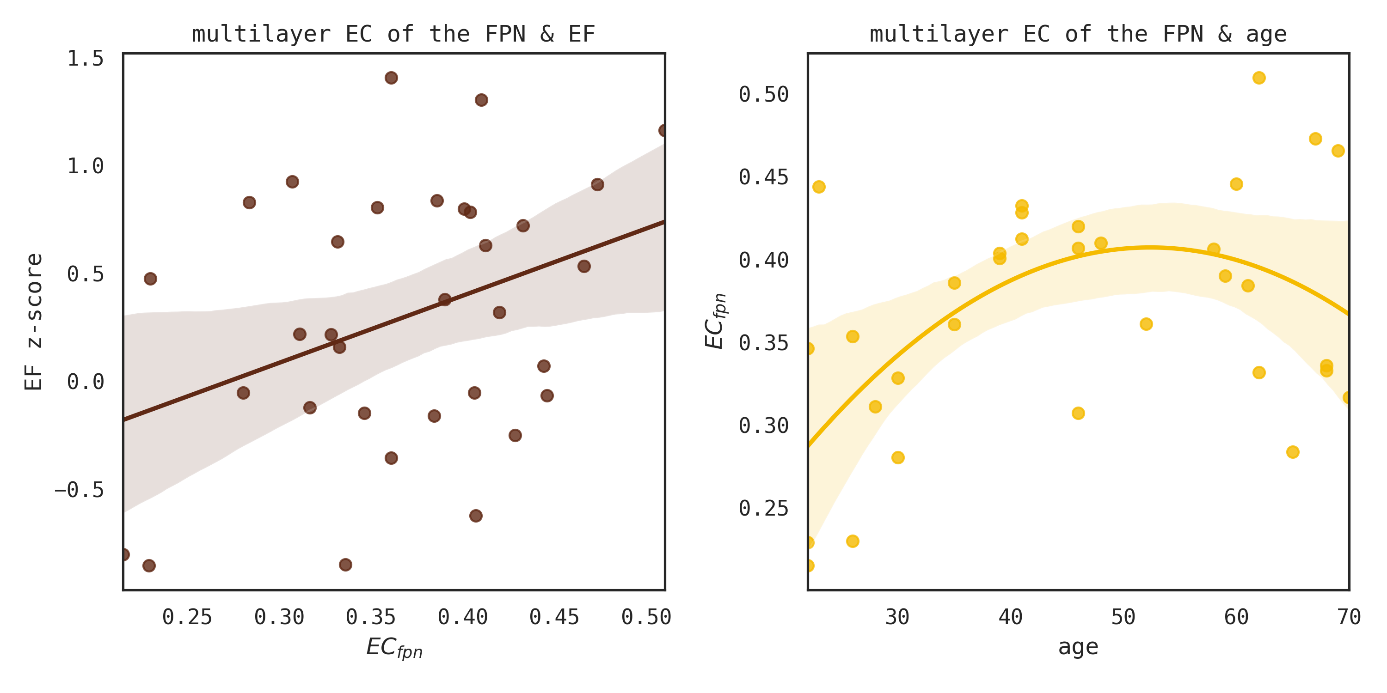

| Supplementary Table 2 | | |  |  |  |  |  |  |
| --- | --- | --- | --- | --- | --- | --- | --- | --- |
| *Hierarchical multiple regression predicting cognitive performance from single- and multilayer EC of the FPN-- BNA atlas* | | | | | | | | |
|  |  |  | Model 1 | | Model 2 | | Model 3 | |
| Variable |  |  | B | β | B | β | B | β |
| (Constant) | |  | 0.416* |  | 0.443* |  | 0.463* |  |
| Age |  |  | -0.023** | -0.179 | -0.023** | -0.179 | -0.023** | -0.180 |
| Sex |  |  | -0.333** | -0.116 | -0.332* | -0.116 | -0.330* | -0.115 |
| *EC_fpn_* single-layer structural network | | | |  | -1.458 | -0.026 | -1.389 | -0.025 |
| *EC_fpn_* single-layer functional network | | | |  | 0.159 | 0.006 | 0.413 | 0.015 |
| *EC_fpn_* multiplex | |  |  |  |  |  | -0.280 | -0.015 |
| *R^2^* |  |  | 0.045 | | 0.046 | | 0.046 | |
| *F* |  |  | 18.277** | | 9.266** | | 7.426** | |
| ∆*R^2^* |  |  | 0.045 | | 0.001 | | 0.000 | |
| *∆F* |  |  | 18.277** | | 0.289 | | 0.106 | |
| ** p<.05, ** p<.001* | |  |  |  |  |  |  |  |
| *Hierarchical multiple regression predicting cognitive performance from single- and multilayer mean eccentricity-- BNA atlas* | | | | | | | | |
|  |  |  | Model 1 | | Model 2 | | Model 3 | |
| Variable |  |  | B | β | B | β | B | β |
| (Constant) | |  | 0.416* |  | 0.417 |  | -1.261 |  |
| Age |  |  | -0.023** | -0.179 | -0.021** | -0.168 | -0.020** | -0.160 |
| Sex |  |  | -0.333** | -0.116 | -0.318* | -0.111 | -0.326* | -0.113 |
| *Ecc_avg_* single-layer structural network | | | |  | -0.014 | -0.048 | -0.014 | -0.048 |
| *Ecc_avg_* single-layer functional network | | | |  | 0.018 | 0.061 | 0.010 | 0.035 |
| *Ecc_avg_* multiplex | |  |  |  |  |  | 0.162 | 0.066 |
| *R^2^* |  |  | 0.045 | | 0.051 | | 0.054 | |
| *F* |  |  | 18.277** | | 10.365** | | 8.886** | |
| ∆*R^2^* |  |  | 0.045 | | 0.006 | | 0.004 | |
| *∆F* |  |  | 18.277** | | 2.387 | | 2.871 | |
| ** p<.05, ** p<.001* | |  |  |  |  |  |  |  |
| *Hierarchical multiple regression predicting multilayer EC of the FPN from age and age squared-- BNA atlas* | | | | | | | | |
|  |  |  | Model 1 | | Model 2 | |  |  |
| Variable |  |  | B | β | B | β |  |  |
| (Constant) | |  | 0.117** |  | 0.159** |  |  |  |
| Age |  |  | 0.000 | -0.062 | -0.003 | -0.484 |  |  |
| Age squared | |  |  |  | 0.000 | 0.427 |  |  |
| *R^2^* |  |  | 0.062 | | 0.084 | |  |  |
| *F* |  |  | 1.651 | | 1.534 | |  |  |
| ∆*R^2^* |  |  | 0.004 | | 0.003 | |  |  |
| *∆F* |  |  | 0.200 | | 0.235 | |  |  |
| ** p<.05, ** p<.001* | |  |  |  |  |  |  |  |
| Supplementary Table 3 | | |  |  |  |  |  |  |
| *Hierarchical multiple regression predicting cognitive performance from single- and multilayer EC of the FPN-- MuMo dataset* | | | | | | | | |
|  |  |  | Model 1 | | Model 2 | |  |  |
| Variable |  |  | B | β | B | β |  |  |
| (Constant) | |  | -0.316 |  | -0.290 |  |  |  |
| *Ecc_fpn_* single-layer structural network | | | 0.848 | 0.194 | 0.577 | 0.132 |  |  |
| *Ecc_fpn_* single-layer functional network | | | 2.828 | 0.109 | 3.334 | 0.129 |  |  |
| *Ecc_fpn_* multiplex | |  |  |  | 1.370 | 0.136 |  |  |
| *R^2^* |  |  | 0.051 | | -0.012 | |  |  |
| *F* |  |  | 0.803 | | 0.673 | |  |  |
| ∆*R^2^* |  |  | 0.051 | | 0.014 | |  |  |
| *∆F* |  |  | 0.803 | | 0.442 | |  |  |
| ** p<.05, ** p<.001* | |  |  |  |  |  |  |  |
| *Hierarchical multiple regression predicting multilayer EC of the FPN from age and age squared-- MuMo dataset* | | | | | | | | |
|  |  |  | Model 1 | | Model 2 | |  |  |
| Variable |  |  | B | β | B | β |  |  |
| (Constant) | |  | 0.085* |  | -0.009 |  |  |  |
| Age |  |  | 0.000 | 0.028 | 0.005 | 1.279 |  |  |
| Age squared | |  |  |  | 0.000 | -1.263 |  |  |
| *R^2^* |  |  | 0.001 | | 0.031 | |  |  |
| *F* |  |  | 0.024 | | 0.478 | |  |  |
| ∆*R^2^* |  |  | 0.001 | | 0.030 | |  |  |
| *∆F* |  |  | 0.024 | | 0.932 | |  |  |
| ** p<.05, ** p<.001* | |  |  |  |  |  |  |  |
